# Microbial cell-free DNA sequencing of bronchoalveolar lavage fluid improves diagnostic yield and may add clinical utility in immunocompromised patients with severe pneumonia

**DOI:** 10.1101/2025.11.05.25339543

**Authors:** Vijeeth Guggilla, Chiagozie O Pickens, Helen K Donnelly, Anna Pawlowski, Erin Korth, Francisco Martinez, Rebecca K Clepp, GR Scott Budinger, Alexander V Misharin, Nandita R Nadig, Benjamin D Singer, Catherine A Gao, Theresa L Walunas, Richard G Wunderink, The NU SCRIPT Study Investigators

## Abstract

**Background:** Despite sophisticated standard of care (SOC) testing (including bronchoalveolar lavage (BAL) fluid culture and multiplex PCR), the etiology of pneumonia in immunocompromised patients is frequently unknown. Microbial cell-free DNA (mcfDNA) sequencing increases diagnostic yield in plasma but remains understudied in BAL fluid.

**Methods:** This was a single center, retrospective, observational cohort study. Residual BAL fluid collected from immunocompromised, mechanically ventilated patients with suspected pneumonia was sent to Karius® for mcfDNA sequencing. Given the retrospective design, mcfDNA sequencing results were unavailable to clinicians. SOC testing results were compared to Karius® BAL (KT-BAL) test reports that classified identified microorganisms as Category One (always pathogenic), Category Two (usually pathogenic), or Category Three (rarely pathogenic).

**Findings:** 228 BAL fluid samples from 155 patients were analyzed. In pneumonia patients, KT-BAL yielded 18 additional Category One organisms, 138 additional Category Two organisms, and 313 additional Category Three organisms compared to SOC. Organisms missed by SOC and identified by KT-BAL included bacterial pathogens (99 patients), clinically relevant DNA viruses (34 patients), non-*Candida* fungi (11 patients), and parasites (1 patient). Compared to patients with Category One or Two organisms identified by both SOC and KT-BAL (n = 68), patients with organisms identified by KT-BAL alone (n = 87) had significantly more cumulative intubation days (11 [5, 20] days vs. 15 [8, 32] days, p = 0.035).

**Interpretation:** KT-BAL increased diagnostic yield for immunocompromised patients with suspected pneumonia with suggestion of prolonged respiratory failure in patients with pathogens missed by SOC.

## Introduction

Pneumonia is the most common infectious cause of death worldwide and disproportionately impacts immunocompromised people.^1–3^ While guideline-based regimens for empiric antibiotic treatment of patients with pneumonia include common pneumonia pathogens, they may be inadequate for more diverse pathogens that cause pneumonia in immunocompromised patients.^4,5^ To ensure appropriate antibiotics are prescribed in this population, clinicians often perform an extensive diagnostic evaluation that includes several antigen-based tests that detect a single organism in the blood, urine, or bronchoalveolar lavage (BAL) fluid, accompanied by cultures. These tests suffer from a lack of sensitivity and sometimes slow turnaround times. The addition of multiplex PCR may increase diagnostic yield and decrease the time to appropriate antibiotic therapy. However, multiplex PCR can only detect a limited number of microbes, and many panels only detect the most common etiologic agents of pneumonia. Indeed, even with advanced diagnostics, an organism is detected in less than half of hospitalized adults with community acquired pneumonia.^6^

In immunocompromised patients, the limitations of existing diagnostics paired with the risk of infection with an uncommon or opportunistic pathogen often results in both antibiotic overtreatment and undertreatment. Prolonged treatment with broad spectrum antibiotics is associated with multiple harms, including increased mortality after adjustment for confounders.^7,8^ Simultaneously, immunocompromised patients are at risk for infection with uncommon pathogens and pathogens that require prolonged or toxic therapies that limit their empiric use, resulting in delayed treatment and poor outcomes. More sensitive and reliable diagnostics for pneumonia can reduce these harms and improve patient outcomes.

Microbial cell-free DNA (mcfDNA) metagenomic sequencing is a culture-independent tool capable of detecting over 1,000 organisms known to infect humans with high sensitivity. While the diagnostic performance of mcfDNA sequencing in plasma specimens has been described, evidence on the performance of mcfDNA sequencing in BAL fluid is limited.^9–11^ We hypothesized that the addition of BAL fluid mcfDNA sequencing would improve the diagnosis of pneumonia in critically ill, immunocompromised patients and that patients with additional pathogens detected by mcfDNA sequencing would associate with distinct clinical outcomes.^12^ To test this, we performed mcfDNA sequencing of residual BAL fluid samples from immunocompromised patients that were prospectively enrolled in an ongoing observational study.^13^ All patients had respiratory failure requiring mechanical ventilation and underwent a clinically indicated BAL.

## Methods

We report this article according to the STROBE checklist (Appendix A).^14^

### Study design and population

This was a retrospective study of patients enrolled in the Successful Clinical Response In Pneumonia Therapy (SCRIPT) Systems Biology Center, a single-center, prospective, observational cohort study of mechanically ventilated patients with suspected pneumonia hospitalized in the medical intensive care unit (ICU) at Northwestern Memorial Hospital (NMH). This study was approved by the Northwestern University Institutional Review Board (STU00204868), which operates under the principles of the Declaration of Helsinki. Patients’ families or licensed authorized representatives consented to take part in the study. SCRIPT began enrolling participants in June 2018 and continues to actively enroll participants who meet eligibility criteria. All participants in SCRIPT have undergone a clinically-indicated bronchoscopic or non-bronchoscopic BAL and have residual BAL fluid available for research analysis.

For this study, participants enrolled in SCRIPT were included if they met criteria for an immunocompromised host, defined as an individual with acute leukemia, human immunodeficiency virus, immunoglobulin deficiency, lymphoma, multiple myeloma, solid organ transplant, stem cell transplant or regular use of any of the following medications in the six months prior to admission: azathioprine, corticosteroids, cyclosporine, cyclophosphamide, mycophenolate, myelosuppressive chemotherapy, rituximab, tacrolimus. This definition aligns with the criteria put forth in the American Thoracic Society Workshop Report on Immunocompromised Host Pneumonia and minimizes selection bias.^12^ The study size was arrived at through these criteria.

### Clinical adjudication

All participants in SCRIPT have a clinically adjudicated diagnosis of pneumonia or non-pneumonia. Adjudication was performed by a panel of five pulmonary and critical care physicians before mcfDNA testing was performed. A diagnosis of bacterial pneumonia, viral pneumonia, bacterial-viral co-infection pneumonia, fungal pneumonia, or non-pneumonia was assigned independently by two of the physician adjudicators. An adjudication of “non-pneumonia control (NPC)” was assigned if BAL fluid analysis and clinical evaluation was not consistent with pneumonia. The NPC samples from immunocompromised subjects were included in the analysis in order to evaluate which organisms are detected even when clinically apparent pneumonia is not present. Pneumonia episodes were also categorized as community-acquired pneumonia (CAP), hospital-acquired pneumonia (HAP), or ventilator-associated pneumonia (VAP). If a discrepancy in the diagnosis was present, a third adjudicator also performed chart review. Complete details of the adjudication protocol have been published previously.^15^

### Sample acquisition and processing

All clinical BAL fluid samples were analyzed in the clinical laboratory with an automated cell count and differential and standard of care (SOC) microbiological testing (see Definitions). After consent was obtained, residual BAL fluid samples were retrieved from the clinical refrigerator and aliquoted for storage at -80°C in a designated research freezer. All samples from patients that met inclusion criteria and no exclusion criteria and with a minimum volume of 0.25 ml were shipped to Karius® for mcfDNA sequencing per company protocol.

### Data acquisition

Demographic, clinical, and microbiologic variables were extracted from the electronic health record (EHR) via the Northwestern Medicine (NM) Enterprise Data Warehouse (NMEDW). All data extraction was performed by NMEDW power users trained for data collection, and manual chart review was performed to verify the quality of extracted data. In the NMEDW, sex assigned at birth is recorded as either male or female based on self-reporting. Ethnicity and race are also recorded based on self-reporting.

Results of mcfDNA sequencing testing by Karius® were sent electronically to our research team in a standardized clinical report format. As this study was conducted retrospectively using residual, cryopreserved BAL fluid, bedside clinical teams could not be aware of the results of mcfDNA sequencing in real time. For each sample, the BAL fluid mcfDNA sequencing test report included the name of each identified organism, the number of microbial reads found for the organism, and the Karius® designation for the pathogenicity of the organism (see Definitions). In addition to the standard information reported, we also obtained the number of total human reads found in the sample.

### Definitions

Standard of care (SOC) testing: SOC microbiological testing for critically ill, immunocompromised patients with suspected pneumonia at our institution includes blood cultures, BAL fluid cell count and differential, BAL quantitative bacterial cultures, fungal cultures, BAL fluid multiplex PCR (BioFire FilmArray Pneumonia Panel), nasopharyngeal swab or BAL fluid multiplex respiratory pathogen PCR (BioFire Respiratory 2.1 Panel), urinary antigens for *Streptococcus pneumoniae* and *Legionella pneumophila*, *Pneumocystis jirovecii* direct fluorescent antibody testing and PCR, BAL fluid galactomannan antigen testing (Bio-Rad Platelia *Aspergillus* Ag), BAL fluid methicillin-resistant *Staphylococcus aureus* (MRSA) PCR (pre-BioFire Pneumonia panel use), and SARS-CoV-2 testing.^16^ Testing was ordered by the clinical team based on clinical suspicion, and the same testing regimen could not be enforced for everyone (i.e., teams may have chosen not to order certain tests).

Karius® Test Pathogen Categories: The operating characteristics of the Karius® BAL test have recently been submitted for publication.^17^ Karius® categorizes organisms based on their pathogenic potential.^18^ Organisms like *P. jirovecii* and *Aspergillus fumigatus* are classified as Category One (always pathogenic). Organisms like *Pseudomonas aeruginosa* and *Haemophilus influenzae* are classified as Category Two (usually pathogenic).

Organisms like *Actinomyces oris* and *Rothia mucilaginosa* are classified as Category Three (rarely pathogenic). Complete lists of Category One, Two, and Three organisms identified in this study are in Supplementary tables 1, 2, and 3, respectively.

### Outcomes and statistics

RNA virus results were excluded from the analysis of diagnostic concordance as mcfDNA sequencing cannot identify RNA viruses. Since autopsy studies of *Candida* detection in respiratory secretions do not document histologically-confirmed pneumonia, *Candida* spp. results were also excluded from SOC and Karius® reports.^19^ For each sample, identified organisms were designated as SOC alone if they were identified by SOC but not by KT-BAL (SOC+/KT-), SOC + KT-BAL if they were identified by both techniques (SOC+/KT+), and KT-BAL alone if they were identified by KT-BAL but not SOC (SOC-/KT+).

Spearman’s rank correlation coefficient was used to assess the correlation between KT-BAL numbers of microbial reads and SOC culture quantities, as well as the correlation between KT-BAL numbers of human reads and markers of host response (i.e., BAL fluid white blood cell counts and BAL fluid percent neutrophils). Mann-Whitney U tests were performed to compare the number of microbial reads for organisms found in some samples by both techniques and in other samples by only KT-BAL, with correction for multiple comparisons via Benjamini-Hochberg. Mann-Whitney U tests were also performed to assess differences in cumulative intubation days, hospital length of stay, and ICU length of stay between SOC+/KT+ patients and SOC-/KT+ patients for Category One or Two organisms. Fisher’s exact test was used to compare ICU mortality between these groups. Continuous variables were reported as medians with first and third quartiles, and categorical variables were reported as counts and percentages. All analyses were performed in Python 3.13.2. A large portion of the clinical data used in this project is available (and de-identified) on Physionet.^13^

## Results

### Characteristics of the study population

704 patients enrolled in SCRIPT between June 2018 and November 2024 were assessed for eligibility. 547 patients were ineligible as they did not meet the inclusion criteria of being immunocompromised. 230 BAL samples from 157 patients were sent to Karius® for sequencing, and 228 sample results from 155 patients were returned after two samples failed quality control checks after receipt at Karius®. 190 samples from 122 immunocompromised pneumonia patients and 38 samples from 33 immunocompromised non-pneumonia control patients were included in the final analysis (Figure 1). The overall cohort had a median [Q1, Q3] age of 61 [52, 68] years and 90/155 (58%) were male. Complete baseline characteristics of the study population are in

**Figure 1.**
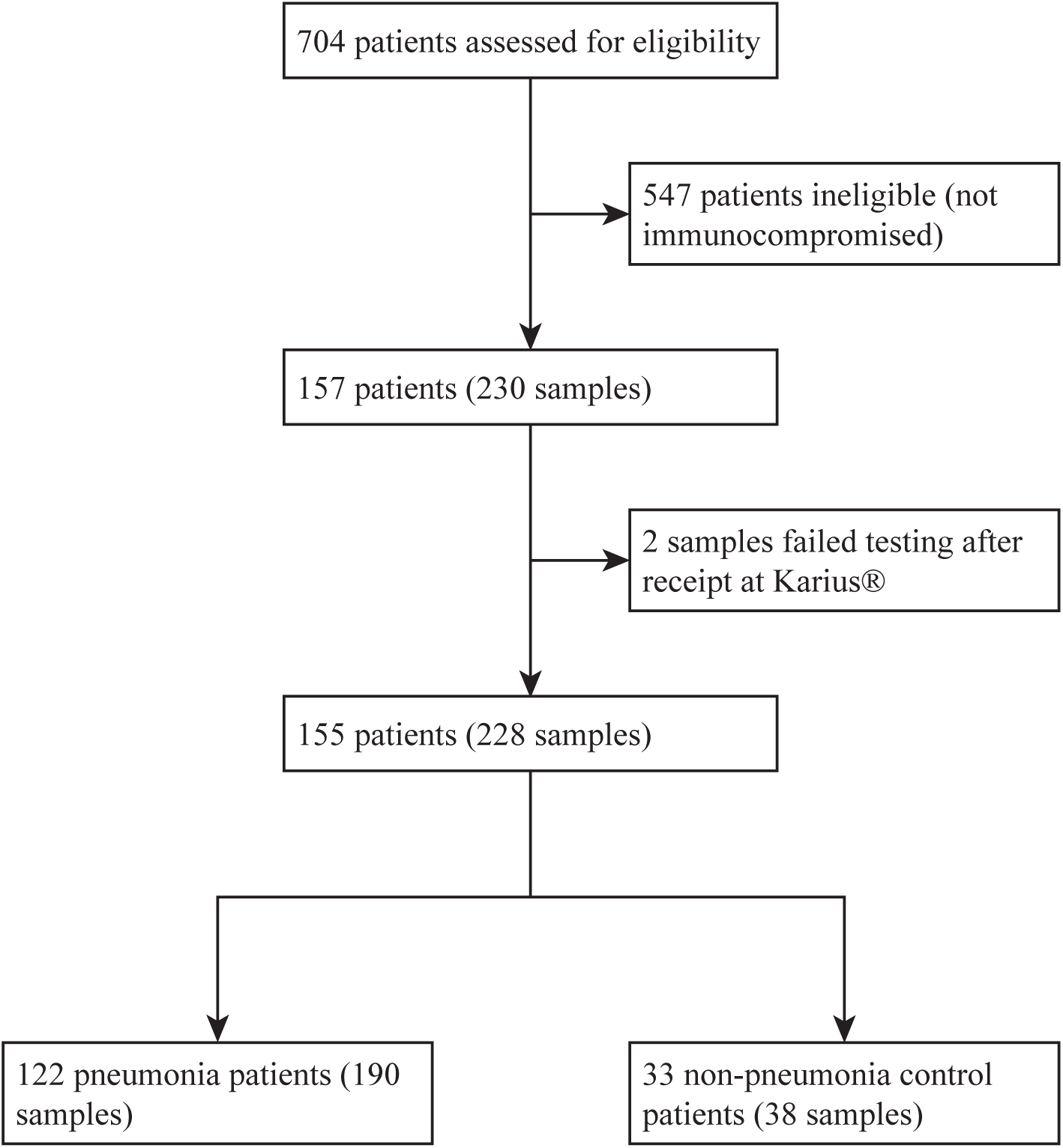
Flow diagram depicting patient and sample selection for the comparison of BAL fluid mcfDNA sequencing with the Karius® test and standard of care testing.

### Additive diagnostic value of KT-BAL

To quantify the additive diagnostic value of KT-BAL, we compared the results of SOC testing and KT-BAL in identifying Category One, Two, and Three organisms across all samples from immunocompromised patients with pneumonia (Figure 2). SOC testing identified five Category One organisms across all samples, and KT- BAL also identified 4/5 (80%) of these organisms, missing *A. fumigatus* in one sample (Supplementary figure 1). KT-BAL also identified an additional 18 organisms across all samples that were missed by SOC testing. This included four samples where KT-BAL identified *A. fumigatus* and four samples where KT-BAL identified *P. jirovecii*. For Category Two, SOC testing identified 111 organisms, and KT-BAL also identified 97/111 (87%) of these organisms. KT-BAL also identified an additional 138 bacterial and viral organisms across all samples that were missed by SOC testing. This included 23 samples where KT-BAL identified Herpes simplex virus type one (HSV-1) and 10 samples where KT-BAL identified *P. aeruginosa*. Finally, for Category Three, KT- BAL identified an additional 313 bacterial organisms across all samples that were not detected by SOC testing.

**Figure 2.**
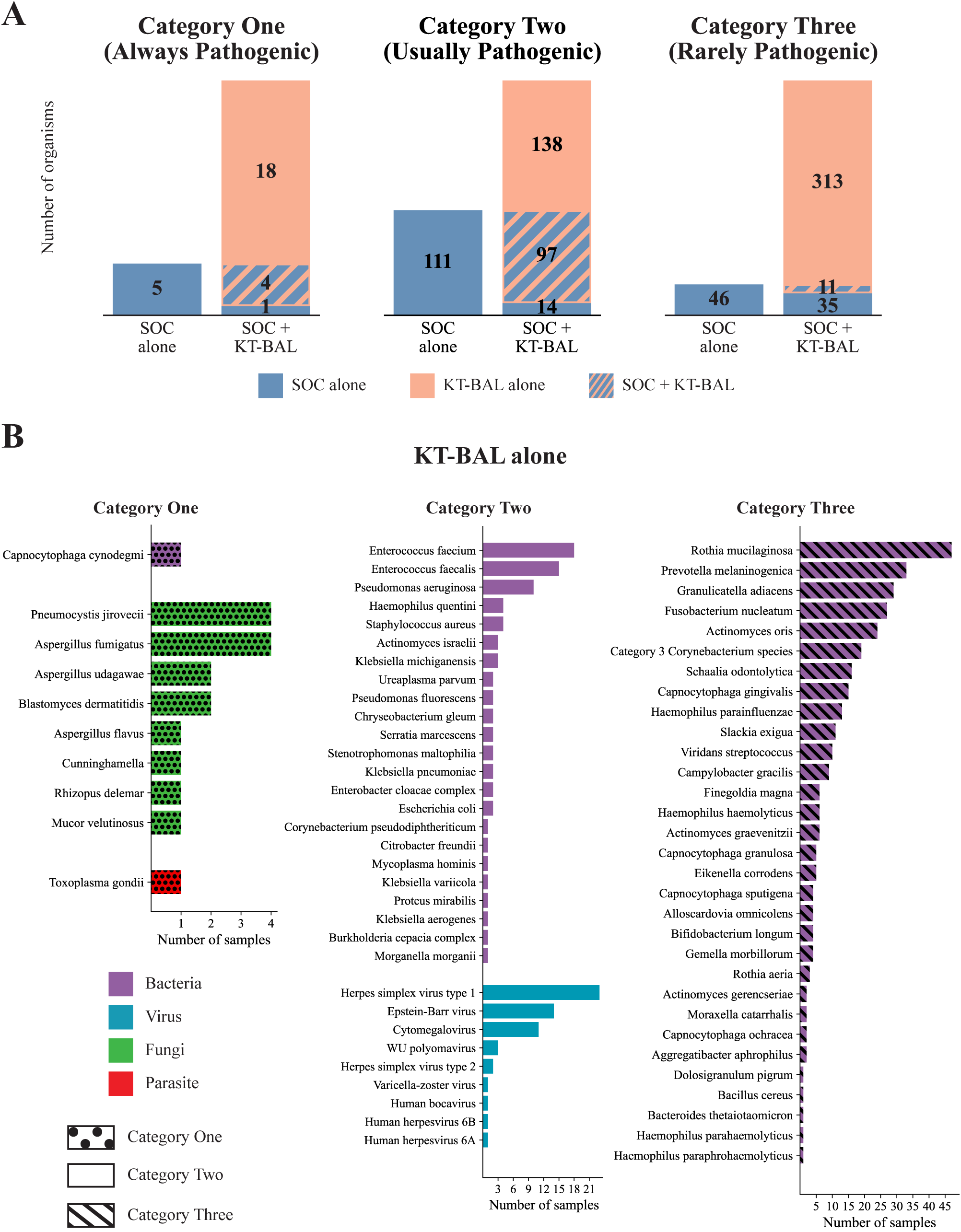
Additive diagnostic value of BAL fluid mcfDNA sequencing with the Karius® test (KT-BAL) compared to standard of care (SOC) testing in immunocompromised patients with pneumonia. A) Number of organisms across all samples identified by either SOC alone, KT-BAL alone, or both techniques, split by KT-BAL category. B) Organisms identified exclusively by KT-BAL across all samples, split by KT-BAL category.

We also performed a sub-analysis examining the additive diagnostic value of KT-BAL in samples from immunocompromised patients who had been adjudicated as microbe-negative pneumonia based on SOC testing not identifying any organisms (Supplementary figure 2). In these samples, KT-BAL identified a variety of bacterial, viral, and fungal organisms, including *P. aeruginosa* in one sample, cytomegalovirus (CMV) in five samples, and *P. jirovecii* in three samples.

In samples from immunocompromised patients adjudicated as NPC, KT-BAL detected Category One or Two pathogens in 17/38 (45%). There were 28 Category One or Two organisms found across these samples, including *P. aeruginosa* in one sample, CMV in four samples, and *P. jirovecii* in three samples, among various other bacterial, viral, fungal, and parasitic organisms (Supplementary figure 3).

**Table 1.** Baseline characteristics of the study population including immunocompromising condition and medication use history.

|  | **Overall** | **Pneumonia patients** | **Non-pneumonia control patients** |
| --- | --- | --- | --- |
|  | n = 155 | n = 122 | n = 33 |
| **Age (years), median [Q1, Q3]** | 61.0 [52.0, 68.0] | 62.5 [53.0, 69.0] | 58.0 [47.0, 65.0] |
| **Gender, n (%)** |  |  |  |
| Male | 90 (58.1) | 76 (62.3) | 14 (42.4) |
| Female | 65 (41.9) | 46 (37.7) | 19 (57.6) |
| **Race, n (%)** |  |  |  |
| Asian | 8 (5.2) | 5 (4.1) | 3 (9.1) |
| Black or African American | 17 (11.0) | 12 (9.8) | 5 (15.2) |
| White | 105 (67.7) | 85 (69.7) | 20 (60.6) |
| Unknown or Not Reported | 25 (16.1) | 20 (16.4) | 5 (15.2) |
| **Ethnicity, n (%)** |  |  |  |
| Hispanic or Latino | 29 (18.7) | 24 (19.7) | 5 (15.2) |
| Not Hispanic or Latino | 119 (76.8) | 93 (76.2) | 26 (78.8) |
| Unknown or Not Reported | 7 (4.5) | 5 (4.1) | 2 (6.1) |
| **BMI, median [Q1, Q3]** | 27.1 [22.8,32.1] | 27.7 [22.8,32.0] | 26.7 [22.8,32.1] |
| **Immune status, n (%)** |  |  |  |
| Immunocompromised | 155 (100.0) | 122 (100.0) | 33 (100.0) |
| **Acute leukemia, n (%)** | 26 (16.8) | 15 (12.3) | 11 (33.3) |
| **HIV, n (%)** | 5 (3.2) | 4 (3.3) | 1 (3.0) |
| **Immunoglobulin deficiency, n (%)** | 10 (6.5) | 8 (6.6) | 2 (6.1) |
| **Lymphoma, n (%)** | 21 (13.5) | 19 (15.6) | 2 (6.1) |
| **Multiple myeloma, n (%)** | 11 (7.1) | 9 (7.4) | 2 (6.1) |
| **Solid organ transplant, n (%)** | 32 (20.6) | 29 (23.8) | 3 (9.1) |
| **Stem cell transplant, n (%)** | 37 (23.9) | 25 (20.5) | 12 (36.4) |
| **Azathioprine, n (%)** | 1 (0.6) | 1 (0.8) | 0 (0.0) |
| **Corticosteroids, n (%)** | 39 (25.2) | 31 (25.4) | 8 (24.2) |
| **Cyclophosphamide, n (%)** | 3 (1.9) | 2 (1.6) | 1 (3.0) |
| **Cyclosporine, n (%)** | 5 (3.2) | 4 (3.3) | 1 (3.0) |
| **Mycophenolate, n (%)** | 38 (24.5) | 30 (24.6) | 8 (24.2) |
| **Myelosuppressive chemotherapy, n (%)** | 30 (19.4) | 20 (16.4) | 10 (30.3) |
| **Rituximab, n (%)** | 16 (10.3) | 15 (12.3) | 1 (3.0) |
| **Tacrolimus, n (%)** | 40 (25.8) | 30 (24.6) | 10 (30.3) |

Abbreviations: BMI, body mass index; HIV: human immunodeficiency virus.

### KT-BAL quantitation of identified organisms

For each organism identified in a sample, the KT-BAL test report includes the number of microbial reads found for that organism. Across all samples, KT-BAL identified organisms present across a wide range of microbial reads (Supplementary figure 4A). Category Three organisms were typically present at lower microbial read levels compared to Category One and Two organisms. Stratifying samples by pneumonia episode type (i.e., NPC, CAP, HAP, VAP) showed that VAP and HAP samples tended to have more Category One and Two organisms present at higher loads compared to NPC and CAP samples (Supplementary figure 4B).

For all Category One or Two organisms that were identified by both techniques in at least five pneumonia samples and identified only by KT-BAL in at least five different pneumonia samples, median [Q1, Q3] microbial load was significantly higher in the congruent samples compared to the KT-BAL only samples (Figure 3A). This included *Enterococcus faecalis* (4052 [2,651, 17,904] reads in congruent vs. 120 [52, 1,528] reads in KT-BAL only, p = 0.034), *E. faecium* (29,542 [2,553, 265,571] reads vs. 101 [43, 618] reads, p = 0.011), and *P. aeruginosa* (13,163 [2,153, 65,808] reads vs. 370 [191, 1,085] reads, p = 0.020).

**Figure 3.**
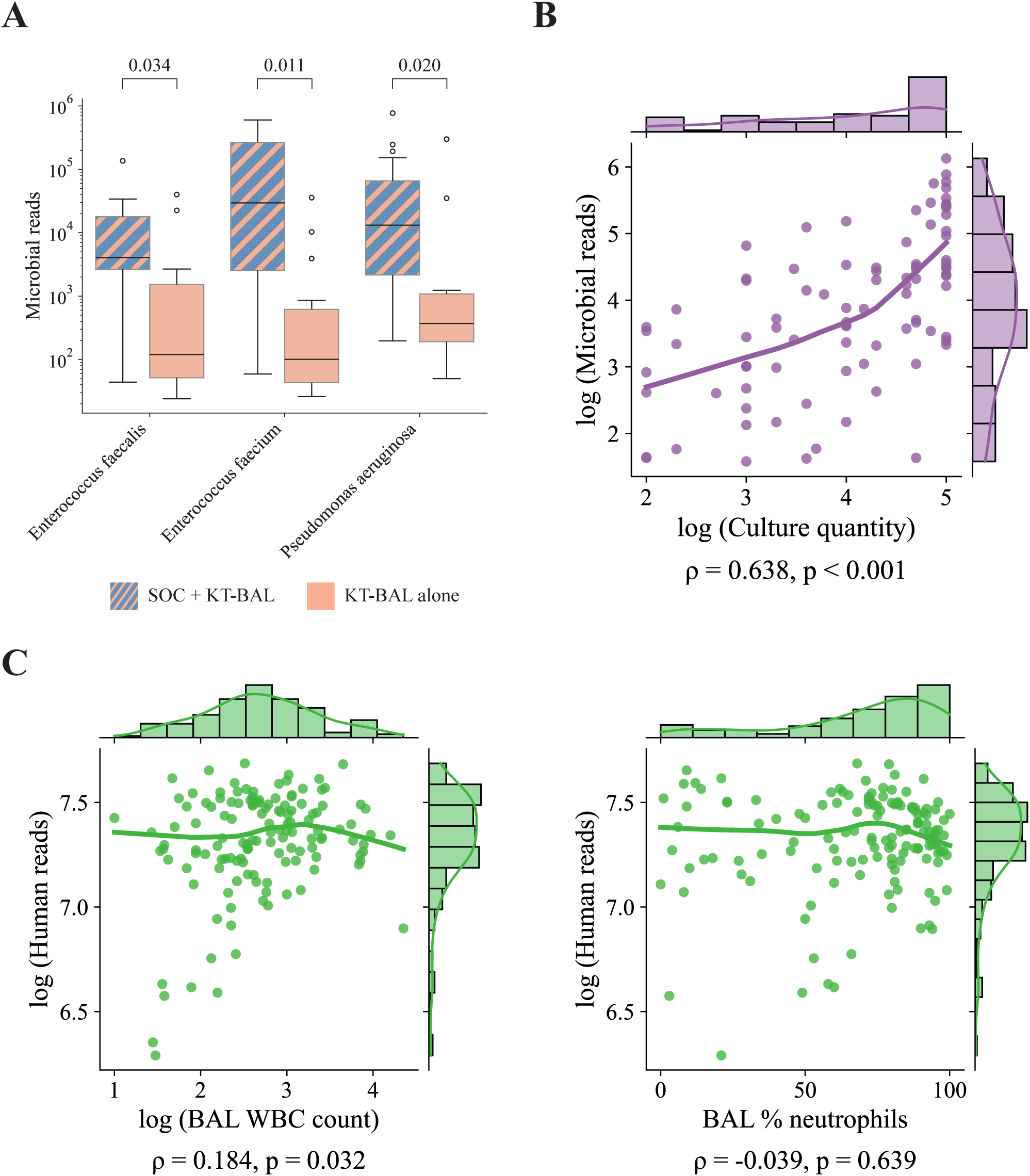
Analysis of microbial read and human read values reported by the BAL fluid Karius® test (KT-BAL). A) Organisms that are identified in at least five samples by both KT-BAL and standard of care (SOC) testing and in at least five other samples by only KT-BAL have significantly lower microbial read values in the SOC + KT-BAL samples (Mann-Whitney U test, FDR < 0.05). B) KT-BAL microbial read values strongly correlate with culture quantities from SOC (Spearman’s rank correlation). C) KT-BAL human read values do not correlate with markers of host immune response (Spearman’s rank correlation).

KT-BAL microbial read values for organisms that were also identified by SOC quantitative culture were moderately correlated with culture quantities (ρ = 0.638, p < 0.001) (Figure 3B). However, KT-BAL human read values showed no correlation with markers of host immune response, including BAL fluid white blood cell counts and BAL fluid percent neutrophils (Figure 3C).

### Clinical outcomes

Across all patients, we examined how clinical outcomes differed for patients in which SOC missed Category One or Two organisms that KT-BAL identified (Figure 4). Compared to patients with concordant detection of Category One or Two organism by SOC and KT-BAL (SOC+/KT+) (n = 68), patients with detection of a Category One or Two organism by KT-BAL alone (SOC-/KT+) (n = 87) had significantly more cumulative intubation days (11 [5, 20] days in SOC+/KT+ vs. 15 [8, 32] days in SOC-/KT+, p = 0.035). While the differences were not statistically significant, the SOC-/KT+ group also had a longer hospital length of stay (29 [18, 43] days in SOC+/KT+ vs. 31 [19, 55] days in SOC-/KT+, p = 0.47), a longer ICU length of stay (18 [10, 30] days in SOC+/KT+ vs. 21 [11, 36] days in SOC-/KT+, p = 0.26), and a higher ICU mortality rate (25/68 (37%) in SOC+/KT+ vs. 46/87 (53%) in SOC-/KT+, p = 0.052).

**Figure 4.**
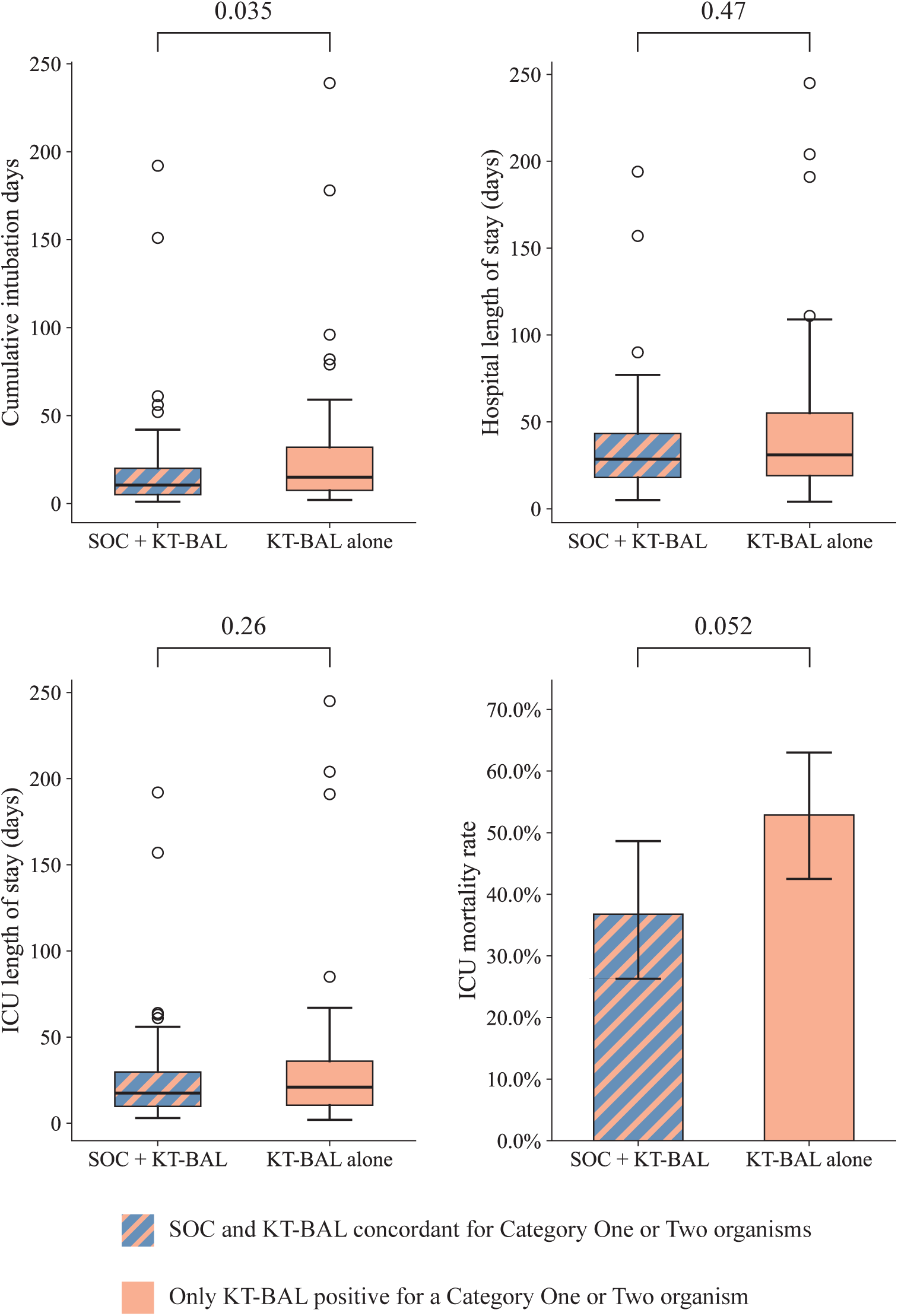
Patients for whom BAL fluid mcfDNA testing with the Karius® test (KT-BAL) identified Category One or Two organisms missed by standard of care (SOC) had significantly more cumulative intubation days compared to patients where KT-BAL and SOC were concordant for all Category One or Two organisms. Hospital length of stay, ICU length of stay, and ICU mortality were worse in the KT-BAL only group, but these differences were not significant (Mann-Whitney U test for continuous variables, Fisher’s exact test for mortality).

## Discussion

In this retrospective cohort study of immunocompromised, mechanically ventilated patients with suspected pneumonia, BAL fluid mcfDNA sequencing with the Karius® test augmented SOC diagnostic testing through increased detection of microbes, including commonly pathogenic organisms (defined as Category One or Category Two organisms by Karius®). In addition, the microbial reads reported by Karius® for these organisms moderately correlated with BAL fluid culture quantities and tended to be higher in nosocomial pneumonia compared to non-pneumonia controls and CAP, demonstrating the increased sensitivity of KT-BAL compared to SOC for pathogen detection.

While others have reported that next-generation sequencing of BAL fluid samples increases the detection of important microbes, the design of our study also allowed us to test associations between molecular detection of an otherwise occult potential pathogen and clinical outcomes.^20–24^ While this study was underpowered to detect differences in some clinical outcomes, patients who had commonly pathogenic organisms exclusively detected by KT-BAL had longer durations of mechanical ventilation compared to patients who had these organisms detected by both KT-BAL and SOC. Taken together, our findings suggest that pneumonia caused by pathogens that go undetected by SOC may lead to missed treatment and be associated with worse clinical outcomes in immunocompromised patients. These associations are clinically plausible as longer duration of intubation may reflect prolonged respiratory failure from persistent lung injury due to an untreated pathogen. For example, *A. fumigatus* and *P. jirovecii* are not covered by broad spectrum anti-bacterial regimens and are well known causes of severe respiratory failure in immunocompromised individuals. Alternatively, molecular detection of a potential pathogen like CMV may be associated with poor host resilience and prolonged respiratory failure, even if the virus is not the agent of active infection. Reactivation of latent pathogens, including CMV, Herpes viruses, and *Pneumocystis*, as a marker of immune status may explain the detections in our patients adjudicated as NPCs. Others have also documented worse outcomes in patients with severe pneumonia and *P. jirovecii* colonization detected by metagenomic sequencing.^25^ Both hypotheses should be prospectively examined to further elucidate the therapeutic and prognostic implications of detecting Category One or Two organisms in BAL fluid.

The microbial reads reported by Karius® provide insights into the limitations of clinical testing. When the same organism was detected in the BAL fluid by both mcfDNA testing and conventional testing with semi-quantitative culture and multiplex PCR, the number of microbial reads detected by mcfDNA sequencing was higher than when only detected by mcfDNA sequencing. This finding reflects the improved sensitivity of molecular tests and demonstrates the limitations of culture-based detection when bacterial burden is low.

Importantly, the minimum inoculum size (indirectly measured by microbial reads, quantitative PCR, or colony forming units) needed to cause pneumonia is unknown. While cfu/mL thresholds of 10^4^ are commonly used to define a BAL culture as positive, infection can occur at lower levels, and colonization without infection may be present at higher values. Our findings also suggest that microbial loads sufficient to cause infection may vary based on pneumonia category, as microbial reads detected by KT-BAL tended to be higher in HAP and VAP compared to CAP. The association between increasing bacterial burden and decreasing diversity has been best documented for VAP. Lower concentrations of newly introduced highly virulent pathogens are an alternative theory of pathogenesis for CAP. The number of microbial reads detected by mcfDNA in BAL fluid provides an additional tool to explore these questions.

We focused our analysis on Category One and Two microbes per the clinically available Karius® reports. Category Three organisms are potentially pathogens in specific circumstances, including immunocompromised patients and patients with large volume aspiration of oropharyngeal secretions.^26,27^ Distinguishing between components of the normal lung microbiome, colonization of central airways, and actual pneumonia is difficult in these cases. Frequent detection of Category Three microbes in our cases independently adjudicated as non-pneumonia controls illustrates this issue.

In addition to routine measures of the immune response (e.g., fever, leukocytosis), BAL neutrophilia is a marker of acute lung injury.^28^ We did not find a correlation between BAL neutrophilia and microbial reads reported by Karius®, further supporting the hypothesis that the minimum microbial burden required for infection likely varies based on the pathogen, category of pneumonia (i.e., CAP, HAP, VAP), and host immune status.

While our study has several strengths, there are also limitations. The retrospective study design limits our control of bias and confounders that may be present when associating KT-BAL detection of a potential pathogen with cumulative duration of intubation. Given the potential clinical implications of this association, prospective studies that minimize the risk of bias are warranted. A second limitation of this study is our enrolled population. All patients were immunocompromised, intubated, mechanically ventilated, and underwent clinical BAL for suspected pneumonia. The findings from our study may not be generalizable to non-intubated or immunocompetent individuals, who are likely to be significantly less ill and have a lower pathogen burden.

Another limitation is our inability to compare the yield of mcfDNA sequencing from matched plasma samples. Existing studies suggest plasma mcfDNA sequencing has high sensitivity for the detection of potential etiologies of pneumonia compared to usual care, but additive diagnostic value of BAL fluid mcfDNA sequencing is unknown. We also did not compare mcfDNA sequencing with shotgun metagenomic sequencing, which is performed on intact cells. Shotgun metagenomic sequencing has been evaluated as a diagnostic tool for pneumonia but is not commercially available and has variable sensitivity and generalizability given non-standardized laboratory protocols and sequencing pipelines when applied to respiratory samples.^29,30^ Finally, this was a single-site study, and the patient population and care delivery approaches at NMH may not be representative of the general patient population. Despite these limitations, our findings support an adequately powered study to prospectively evaluate the clinical impact of BAL fluid mcfDNA testing in immunocompromised patients with known or suspected pneumonia.

## Data Availability

Individual participant data cannot be shared publicly because the data contain protected health information (including identifying or sensitive patient information). Data are available from the corresponding author upon reasonable request for researchers who meet the criteria for access to confidential data and sign a data use agreement. In addition, a large portion of the clinical data collected from the participants in the SCRIPT study has been deidentified and is available on PhysioNet to those who complete the outlined credentialing and training.

https://doi.org/10.13026/w7kd-qj96

## Declaration of generative AI and AI-assisted technologies in the manuscript preparation process

During the preparation of this work the authors used Claude to help with coding. After using this tool/service, the authors reviewed and edited the content as needed and take full responsibility for the content of the published article.

## Data sharing

Individual participant data cannot be shared publicly because the data contain protected health information (including identifying or sensitive patient information). Data are available from the corresponding author upon reasonable request for researchers who meet the criteria for access to confidential data and sign a data use agreement. In addition, a large portion of the clinical data collected from the participants in the SCRIPT study has been deidentified and is available on PhysioNet to those who complete the outlined credentialing and training.^13^

## Contributors’ statement

Vijeeth Guggilla (Data curation, Formal analysis, Investigation, Methodology, Software, Visualization, Writing original draft, Writing - review & editing), Chiagozie O Pickens (Conceptualization, Formal analysis, Investigation, Methodology, Supervision, Validation, Writing - original draft, Writing - review & editing), Helen K Donnelly (Data curation, Investigation, Project administration, Writing - review & editing), Anna Pawlowski (Data curation, Investigation, Writing - review & editing), Erin Korth (Investigation, Writing - review & editing), Francisco Martinez (Investigation, Writing - review & editing), Rebecca K Clepp (Data curation, Investigation, Project administration, Writing - review & editing), GR Scott Budinger (Conceptualization, Methodology, Resources, Writing - review & editing), Alexander V Misharin (Conceptualization, Formal analysis, Methodology, Visualization, Writing - review & editing), Nandita R Nadig (Conceptualization, Writing - review & editing), Benjamin D Singer (Conceptualization, Writing - review & editing), Catherine A Gao (Conceptualization, Data curation, Methodology, Supervision, Visualization, Writing - review & editing), Theresa L Walunas (Resources, Supervision, Writing - review & editing), and Richard G Wunderink (Conceptualization, Formal analysis, Funding acquisition, Methodology, Project administration, Resources, Validation, Writing - original draft, Writing - review & editing)

