## Supplementary material for "Microbial cell-free DNA sequencing of bronchoalveolar lavage fluid improves diagnostic yield and may add clinical utility in immunocompromised patients with severe pneumonia"

**Table of contents**

*Supplementary tables*

Supplementary table 1. ---------- 2

Supplementary table 2. ---------- 3

Supplementary table 3. ---------- 4

*Supplementary figures*

Supplementary figure 1. ---------- 5

Supplementary figure 2. ---------- 6

Supplementary figure 3. ---------- 7

Supplementary figure 4. ---------- 8

*Appendices*

Appendix A. ---------- 9-11

**Supplementary table 1.** All unique organisms identified by BAL fluid mcfDNA sequencing with the Karius® test across all samples in the study designated as Category One organisms by Karius®.

| **Organism name** | **Species type** | **Category** |
|---|---|---|
| *Capnocytophaga cynodegmi* | Bacteria | Category One |
| *Legionella pneumophila* | Bacteria | Category One |
| *Nocardia cyriacigeorgica* | Bacteria | Category One |
| *Tropheryma whipplei* | Bacteria | Category One |
| *Aspergillus flavus* | Fungi | Category One |
| *Aspergillus fumigatus* | Fungi | Category One |
| *Aspergillus udagawae* | Fungi | Category One |
| *Blastomyces dermatitidis* | Fungi | Category One |
| *Cunninghamella* | Fungi | Category One |
| *Mucor velutinosus* | Fungi | Category One |
| *Pneumocystis jirovecii* | Fungi | Category One |
| *Rhizomucor pusillus* | Fungi | Category One |
| *Rhizopus delemar* | Fungi | Category One |
| *Toxoplasma gondii* | Parasite | Category One |

**Supplementary table 2.** All unique organisms identified by BAL fluid mcfDNA sequencing with the Karius® test across all samples in the study designated as Category Two organisms by Karius®.

| Organism name | Species type | Category |
|---|---|---|
| *Achromobacter xylosoxidans* | Bacteria | Category Two |
| *Acinetobacter baumannii* | Bacteria | Category Two |
| *Actinomyces israelii* | Bacteria | Category Two |
| *Burkholderia cepacia complex* | Bacteria | Category Two |
| *Chryseobacterium gleum* | Bacteria | Category Two |
| *Citrobacter freundii* | Bacteria | Category Two |
| *Corynebacterium pseudodiphtheriticum* | Bacteria | Category Two |
| *Elizabethkingia anophelis* | Bacteria | Category Two |
| *Enterobacter cloacae complex* | Bacteria | Category Two |
| *Enterococcus faecalis* | Bacteria | Category Two |
| *Enterococcus faecium* | Bacteria | Category Two |
| *Escherichia coli* | Bacteria | Category Two |
| *Haemophilus influenzae* | Bacteria | Category Two |
| *Haemophilus quentini* | Bacteria | Category Two |
| *Klebsiella aerogenes* | Bacteria | Category Two |
| *Klebsiella michiganensis* | Bacteria | Category Two |
| *Klebsiella oxytoca* | Bacteria | Category Two |
| *Klebsiella pneumoniae* | Bacteria | Category Two |
| *Klebsiella variicola* | Bacteria | Category Two |
| *Morganella morganii* | Bacteria | Category Two |
| *Mycoplasma hominis* | Bacteria | Category Two |
| *Proteus mirabilis* | Bacteria | Category Two |
| *Pseudomonas aeruginosa* | Bacteria | Category Two |
| *Pseudomonas fluorescens* | Bacteria | Category Two |
| *Serratia marcescens* | Bacteria | Category Two |
| *Staphylococcus aureus* | Bacteria | Category Two |
| *Stenotrophomonas maltophilia* | Bacteria | Category Two |
| *Streptococcus agalactiae* | Bacteria | Category Two |
| *Ureaplasma parvum* | Bacteria | Category Two |
| Cytomegalovirus | Virus | Category Two |
| Epstein-Barr virus | Virus | Category Two |
| Herpes simplex virus type 1 | Virus | Category Two |
| Herpes simplex virus type 2 | Virus | Category Two |
| Human bocavirus | Virus | Category Two |
| Human herpesvirus 6A | Virus | Category Two |
| Human herpesvirus 6B | Virus | Category Two |
| Varicella-zoster virus | Virus | Category Two |
| WU polyomavirus | Virus | Category Two |

**Supplementary table 3.** All unique organisms identified by BAL fluid mcfDNA sequencing with the Karius® test across all samples in the study designated as Category Three organisms by Karius®.

| **Organism name** | **Species type** | **Category** |
|---|---|---|
| *Actinomyces gerencseriae* | Bacteria | Category Three |
| *Actinomyces graevenitzii* | Bacteria | Category Three |
| *Actinomyces oris* | Bacteria | Category Three |
| *Aggregatibacter aphrophilus* | Bacteria | Category Three |
| *Alloscardovia omnicolens* | Bacteria | Category Three |
| *Bacillus cereus* | Bacteria | Category Three |
| *Bacteroides thetaiotaomicron* | Bacteria | Category Three |
| *Bifidobacterium longum* | Bacteria | Category Three |
| *Campylobacter gracilis* | Bacteria | Category Three |
| *Capnocytophaga gingivalis* | Bacteria | Category Three |
| *Capnocytophaga granulosa* | Bacteria | Category Three |
| *Capnocytophaga ochracea* | Bacteria | Category Three |
| *Capnocytophaga sputigena* | Bacteria | Category Three |
| *Category 3 Corynebacterium species* | Bacteria | Category Three |
| *Dolosigranulum pigrum* | Bacteria | Category Three |
| *Eikenella corrodens* | Bacteria | Category Three |
| *Finegoldia magna* | Bacteria | Category Three |
| *Fusobacterium necrophorum* | Bacteria | Category Three |
| *Fusobacterium nucleatum* | Bacteria | Category Three |
| *Gemella morbillorum* | Bacteria | Category Three |
| *Granulicatella adiacens* | Bacteria | Category Three |
| *Haemophilus haemolyticus* | Bacteria | Category Three |
| *Haemophilus parahaemolyticus* | Bacteria | Category Three |
| *Haemophilus parainfluenzae* | Bacteria | Category Three |
| *Haemophilus paraphrohaemolyticus* | Bacteria | Category Three |
| *Moraxella catarrhalis* | Bacteria | Category Three |
| *Prevotella melaninogenica* | Bacteria | Category Three |
| *Rothia aeria* | Bacteria | Category Three |
| *Rothia mucilaginosa* | Bacteria | Category Three |
| *Schaalia odontolytica* | Bacteria | Category Three |
| *Slackia exigua* | Bacteria | Category Three |
| *Viridans streptococcus* | Bacteria | Category Three |

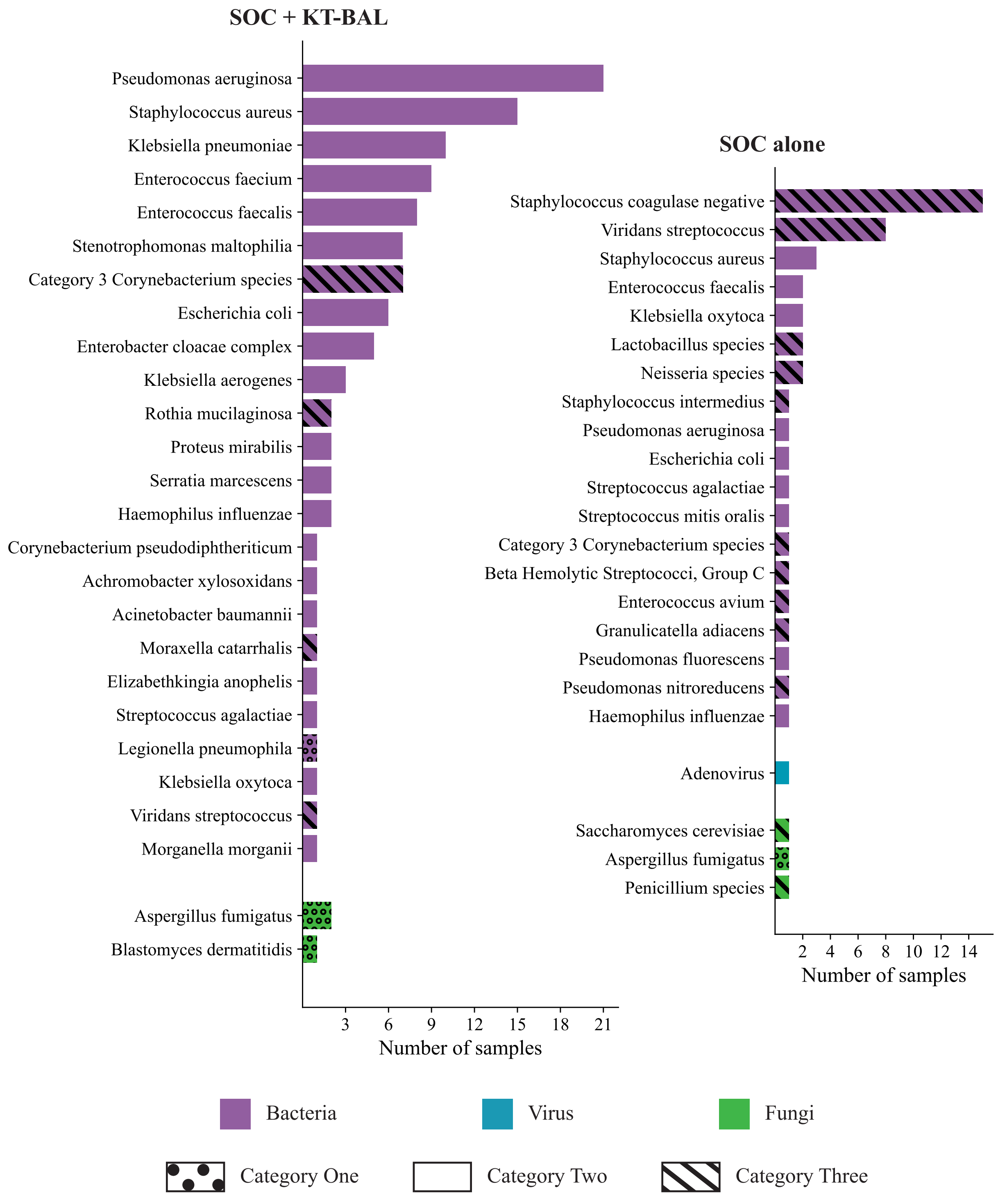


**Supplementary figure 1.** Organisms identified by both standard of care (SOC) and BAL fluid mcfDNA sequencing with the Karius® test (KT-BAL) and organisms exclusively identified by SOC across all samples from immunocompromised patients with pneumonia.

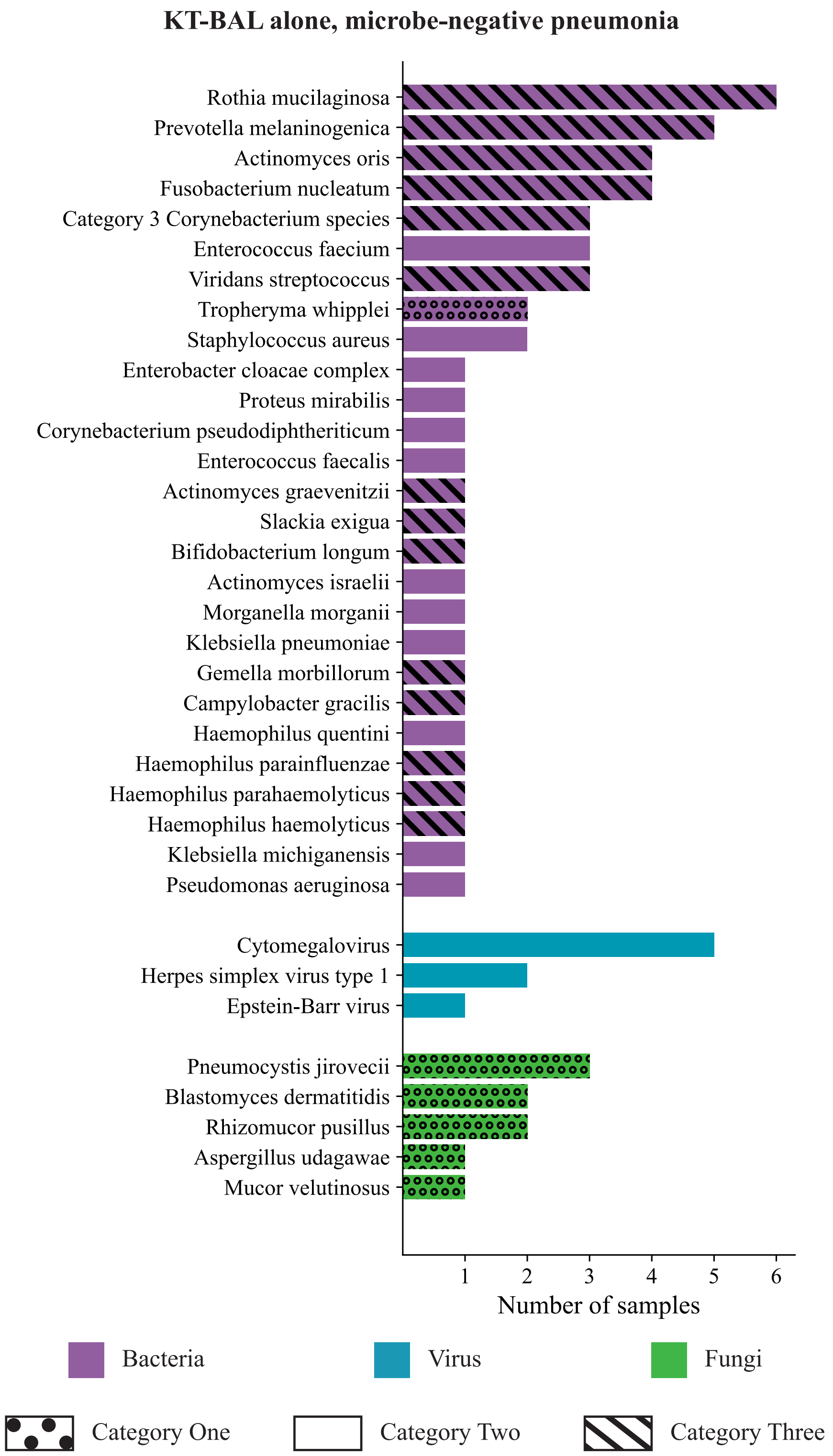


**Supplementary figure 2.** Organisms exclusively identified by BAL fluid mcfDNA sequencing with the Karius® test (KT-BAL) across all samples from immunocompromised patients with microbe-negative pneumonia based on standard of care testing not identifying any organisms.

**KT-BAL alone, non-pneumonia controls**

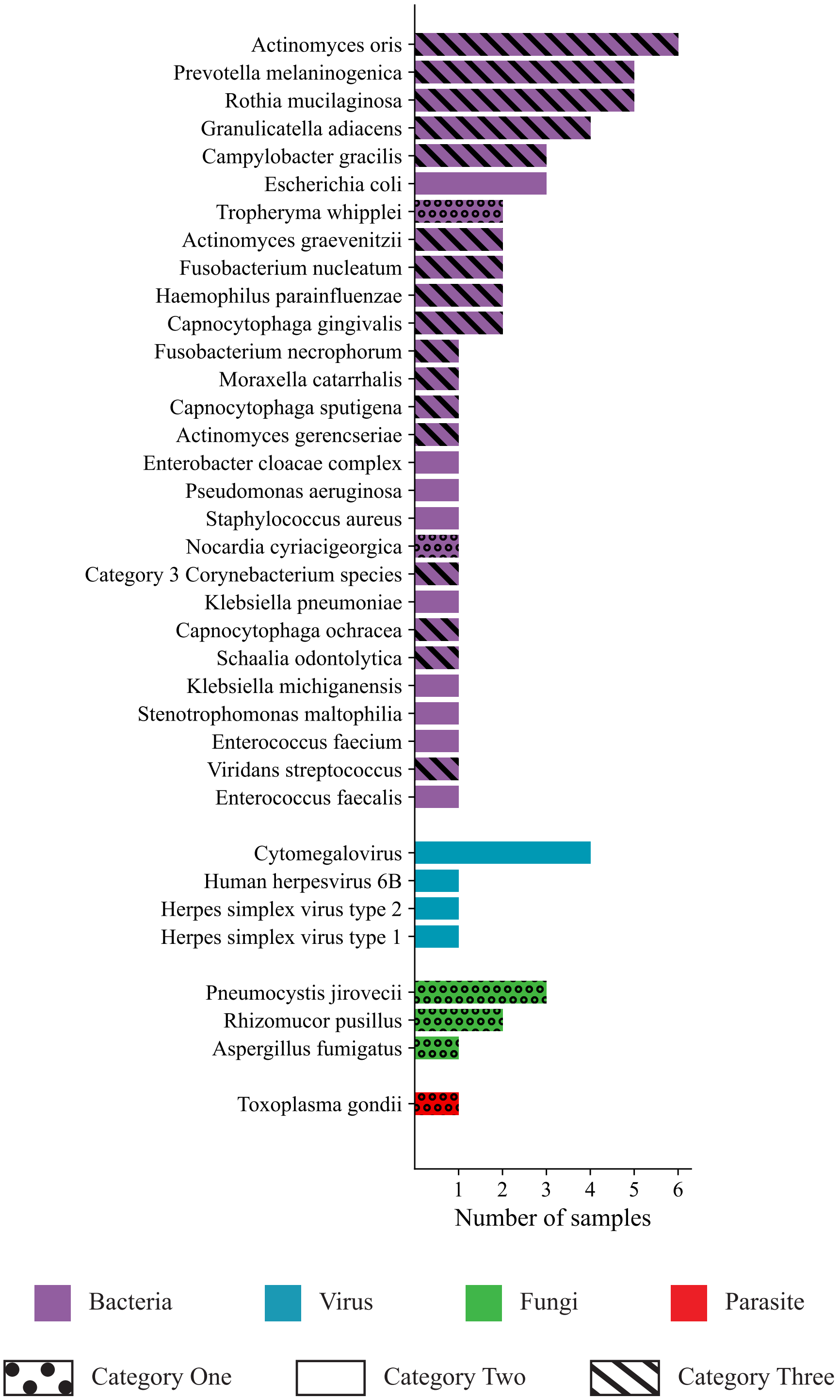


**Supplementary figure 3.** Organisms exclusively identified by BAL fluid mcfDNA sequencing with the Karius® test (KT-BAL) across all samples from immunocompromised non-pneumonia control (NPC) patients.

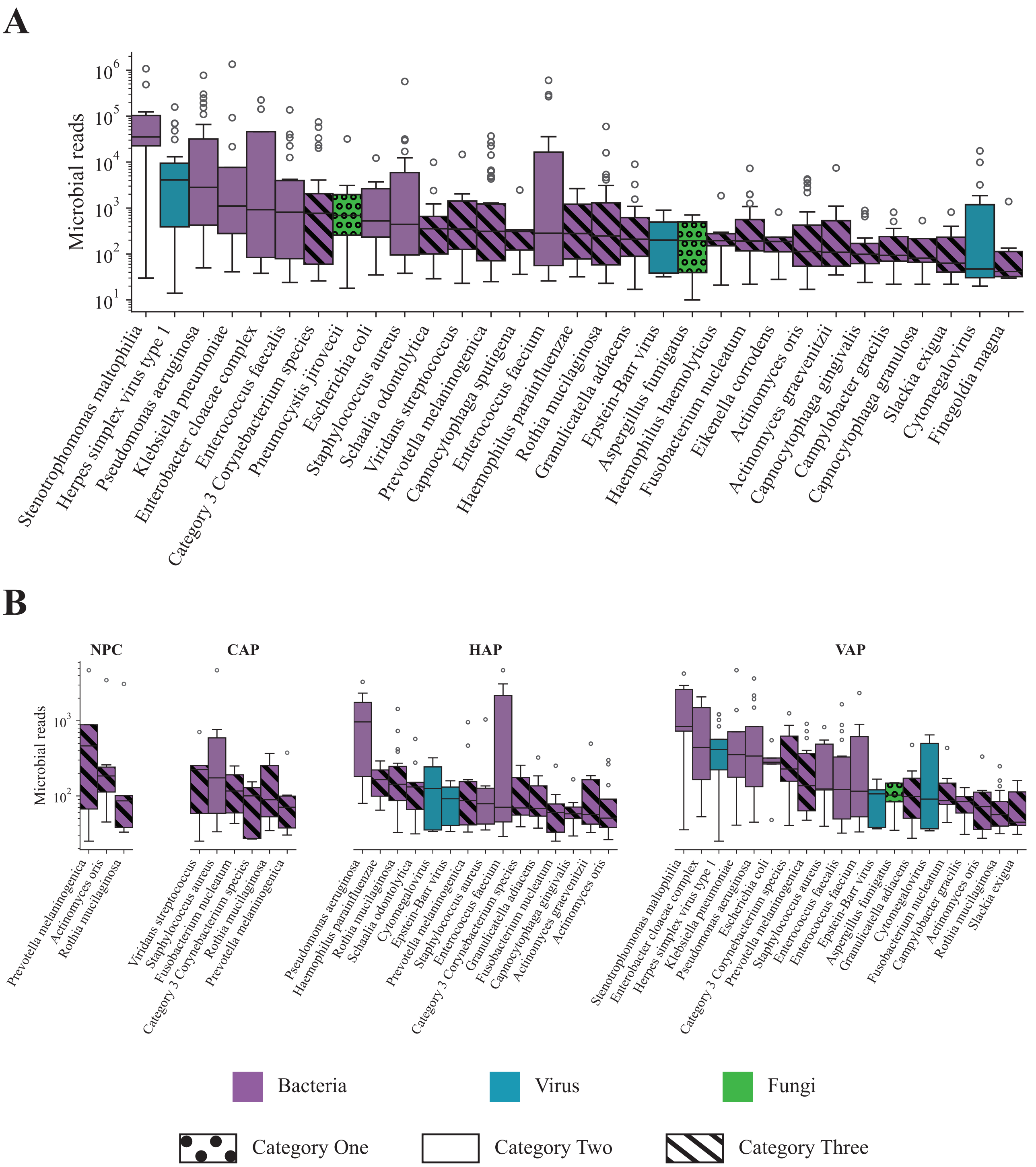


**Supplementary figure 4.** A) Median microbial read values of all organisms identified by BAL fluid mcfDNA sequencing with the Karius® test (KT-BAL) in at least five samples. B) Median microbial read values of all organisms identified by KT-BAL in at least five samples of a given episode type, split by episode type: non-pneumonia control (NPC), community-acquired pneumonia (CAP), hospital-acquired pneumonia (HAP), and ventilator-associated pneumonia (VAP).

**Appendix A.** STROBE checklist of items that should be included in reports of cohort studies.

| | Item No | Recommendation |
|---|---|---|
| **Title and abstract** | 1 | (*a*) Indicate the study's design with a commonly used term in the title or the abstract<br>**Page 2** |
| | | (*b*) Provide in the abstract an informative and balanced summary of what was done and what was found<br>**Page 2** |
| **Introduction** | | |
| Background/rationale | 2 | Explain the scientific background and rationale for the investigation being reported<br>**Page 3** |
| Objectives | 3 | State specific objectives, including any prespecified hypotheses<br>**Page 3-4** |
| **Methods** | | |
| Study design | 4 | Present key elements of study design early in the paper<br>**Page 4** |
| Setting | 5 | Describe the setting, locations, and relevant dates, including periods of recruitment, exposure, follow-up, and data collection<br>**Page 4** |
| Participants | 6 | (*a*) Give the eligibility criteria, and the sources and methods of selection of participants. Describe methods of follow-up<br>**Page 4** |
| | | (*b*) For matched studies, give matching criteria and number of exposed and unexposed<br>**N/A** |
| Variables | 7 | Clearly define all outcomes, exposures, predictors, potential confounders, and effect modifiers. Give diagnostic criteria, if applicable<br>**Page 4, 7** |
| Data sources/ measurement | 8* | For each variable of interest, give sources of data and details of methods of assessment (measurement). Describe comparability of assessment methods if there is more than one group<br>**Page 5-6** |
| Bias | 9 | Describe any efforts to address potential sources of bias<br>**Page 4** |
| Study size | 10 | Explain how the study size was arrived at<br>**Page 4** |
| Quantitative variables | 11 | Explain how quantitative variables were handled in the analyses. If applicable, describe which |

| | | |
|---|---|---|
| | | groupings were chosen and why<br>**Page 7** |
| Statistical methods | 12 | (*a*) Describe all statistical methods, including those used to control for confounding<br>**Page 7** |
| | | (*b*) Describe any methods used to examine subgroups and interactions<br>**Page 6-7** |
| | | (*c*) Explain how missing data were addressed<br>**N/A** |
| | | (*d*) If applicable, explain how loss to follow-up was addressed<br>**N/A** |
| | | (*e*) Describe any sensitivity analyses<br>**N/A** |
| **Results** | | |
| Participants | 13* | (a) Report numbers of individuals at each stage of study—eg numbers potentially eligible, examined for eligibility, confirmed eligible, included in the study, completing follow-up, and analysed<br>**Page 7-8** |
| | | (b) Give reasons for non-participation at each stage<br>**Page 7-8** |
| | | (c) Consider use of a flow diagram<br>**Figure 1** |
| Descriptive data | 14* | (a) Give characteristics of study participants (eg demographic, clinical, social) and information on exposures and potential confounders<br>**Page 8, Table 1** |
| | | (b) Indicate number of participants with missing data for each variable of interest<br>**N/A** |
| | | (c) Summarise follow-up time (eg, average and total amount)<br>**N/A** |
| Outcome data | 15* | Report numbers of outcome events or summary measures over time<br>**Page 12, 15** |
| Main results | 16 | (*a*) Give unadjusted estimates and, if applicable, confounder-adjusted estimates and their precision (eg, 95% confidence interval). Make clear which confounders were adjusted for and why they were included<br>**Page 8, 12, 15** |

| | | |
|---|---|---|
| | | (*b*) Report category boundaries when continuous variables were categorized<br><br>**N/A** |
| | | (*c*) If relevant, consider translating estimates of relative risk into absolute risk for a meaningful time period<br><br>**N/A** |
| Other analyses | 17 | Report other analyses done—eg analyses of subgroups and interactions, and sensitivity analyses<br><br>**Page 8, 12** |
| **Discussion** | | |
| Key results | 18 | Summarise key results with reference to study objectives<br><br>**Page 15-16** |
| Limitations | 19 | Discuss limitations of the study, taking into account sources of potential bias or imprecision. Discuss both direction and magnitude of any potential bias.<br><br>**Page 17** |
| Interpretation | 20 | Give a cautious overall interpretation of results considering objectives, limitations, multiplicity of analyses, results from similar studies, and other relevant evidence<br><br>**Page 15-17** |
| Generalisability | 21 | Discuss the generalisability (external validity) of the study results<br><br>**Page 17** |
| **Other information** | | |
| Funding | 22 | Give the source of funding and the role of the funders for the present study and, if applicable, for the original study on which the present article is based<br><br>**Page 7, 18** |

*Give information separately for exposed and unexposed groups.
